# Identifying Prepubertal Children with Risk for Suicide Using Deep Neural Network Trained on Multimodal Brain Imaging-Derived Phenotypes

**DOI:** 10.1101/2021.10.10.21264580

**Authors:** Bo-Gyeom Kim, Gun Ahn, Sooyoung Kim, Kakyeong Kim, Hyeonjin Kim, Eunji Lee, Woo-Young Ahn, Jae-Won Kim, Jiook Cha

## Abstract

Suicide is among the leading causes of death in youth worldwide. Early identification of children with high risk for suicide is key to effective screening and prevention strategies. Brain imaging can show functional or structural abnormalities related to youth suicidality, but literature is scarce. Here we tested the extent to which brain imaging is useful in predicting suicidal risk in children. In the largest to date, multi-site, multi-ethnic, epidemiological developmental samples in the US (N = 6,172; the ABCD study), we trained and validated machine learning models and deep neural networks on the multimodal brain imaging derived phenotypes (morphometry, white matter connectivity, functional activation, and connectivity) along with behavioral and self-reported psychological questionnaire data. The model trained on diffusion white matter connectomes showed the best performance (test AUC-ROC = 74.82) with a one percentage increase compared with the baseline model trained on behavioral and psychological data (test AUC-ROC = 74.16). Models trained on other MRI modalities showed similar but slightly lower performances. Model interpretation showed the important brain features involved in attention, emotion regulation, and motor coordination, such as the anterior cingulate cortex, temporal gyrus, and precentral gyrus. It further showed that the interaction of brain features with depression and impulsivity measures contributed to the optimal prediction of youth suicidality. This study demonstrates the potential utility of a multimodal brain imaging approach to youth suicidality prediction and uncovers the relationships of the psychological and multi-dimensional and multi-modal neural features to youth suicidality.

## Introduction

Suicide is a major public health problem, with more than 25 million suicide attempts occurring each year worldwide. In youth, suicide is among the leading causes of death. Most suicide research focuses on the adult population ^1^, and the literature on youth suicidality has been scarce. Identifying the neural underpinnings of youth suicidality will advance research by allowing brain-based markers and targets for prediction, monitoring and prevention ^2^. Also, early detection of youth suicidality may not only prevent death by suicide among children but also reduce the risk of psychopathology later in life.

Prior brain imaging literature suggests a link between youth suicidality and abnormally delayed neurocognitive development. Children with suicidal ideation showed a reduced brain response to reward ^3^ or decreased cognitive capacity ^4^. As abnormal neurocognitive development may cause long-term sequelae, young children with prior suicide ideation are more likely prone to the risk for suicide in their later life.

Based on recent findings of brain abnormalities linked to suicidality, several studies applied machine learning to brain imaging data in small samples. Gosnell et al. classified suicidal psychiatric inpatients and non-suicidal inpatients with resting-state functional connectivity and structural neuroimaging^5,6^. Just et al. demonstrated that the functional coupling of the brain regions like the superior medial frontal and anterior cingulate regions, during death-related and life-related concepts, could identify youth with suicidal ideation ^7^. Although previous studies show promise, the practical utility of neuroimaging-based prediction of youth suicidality remains untested rigorously in sufficiently large, representative samples using rigorous predictive modeling approaches.

Multimodal data integration may help predict. Combining the brain’s structural and functional aspects, multimodal brain imaging may better account for complex and dimensional brain abnormalities related to suicidality compared with a single modality approach ^8^. No literature in suicidality research, however, has provided information of what combinations of imaging modalities would optimally explain brain abnormalities related to youth suicidality.

Here, we aim to identify youth with suicidal ideation using deep learning trained on the brain structural, functional, and connectivity data, as well as behavioral and psychological measures. Beyond prediction, using interpretable deep learning, we aim to identify brain circuit features and psychological measures of which nonlinear interactions may contribute to prediction. Leveraging large, representative, epidemiological, multi-site, and developmental study in the US, we test the generalizability and practicality of the prediction models. We also test the utility of advanced computational learning strategies, such as stacking ensembles for data integration and GANs for data augmentation in psychiatric research.

## Methods

### Study Sample

The Adolescent Brain and Cognitive Development (ABCD) study recruited a nationally representative cohort of 9- and 10-year-old children from 21 research sites across the US ^4^. For the current analysis, we used minimally preprocessed data from the curated ABCD annual release 2.0.1 and selected ABCD participants based on the availability of non-imaging measures and neuroimaging data. The number of participants was different in each dataset: sMRI, dMRI, and rs-fMRI dataset included 5,878 participants (783 cases and 5044 controls for suicidal ideation; 48 cases and 5779 controls for suicidal attempt). SST fMRI, N-back fMRI, and MID fMRI included (592 cases and 4035 controls for suicidal ideation; 34 cases and 4596 controls for suicidal attempt). Thus, total participants consisted of 6,172 children (837 cases and 5,335 controls).

### Non-imaging features

#### Clinical Outcomes

Suicidal ideation and attempts were assessed from the computerized version of Kiddie Schedule for Affective Disorders and Schizophrenia (KSADS-COMP) reported by children or caregivers ^9^. Validity of the computerized version of KSADS has been shown elsewhere^9^. Suicidal ideation was constructed to measure both passive suicidal ideation and active suicidal ideation^10^. Among the reports from children and parents, we used ones reporting more severe symptoms. In this study, we used suicide ideation as the main outcome and suicide attempt as the secondary target, given the data availability (fewer numbers of participants with suicide attempts than ones with suicide ideation) and the consideration that suicide ideation precedes suicide attempt.

#### Psychosocial features

To estimate the feasibility of a brain-based prediction model, we constructed a psychosocial model as a benchmark model. The benchmark model consisted of demographic, psychological health, mental health, cognition, and environmental variables, which reported associations with suicidality in the literature.

Sociodemographic variables we considered were age, sex, race, highest parental education, family income, and parental marriage status. Physical health variables included anthropometric variables (height, weight, BMI, and brain volume) and sleep function assessed by Sleep Disturbances Scale for Children ^11^. For mental health, we measured dimensional psychopathology (the Child Behavior Checklist ^12^), impulsivity (UPPS-P ^13^), mania symptoms (Parent General Behavior Inventory^14^), psychosis (ABCD prodromal psychosis scales ^15^), and behavioral inhibition/approach system (PhenX modified version ^16^). Cognitive ability was measured using NIH toolbox ^17^. Environmental variables contained the measures of family conflict, early life stress (ELS), and friendship. Family conflict consisted of nine measures, which were assessed by ABCD Youth Family Environment Scale-Family Conflict Subscale Modified from PhenX. The ELS measures were composite variables based on child exposure domains in the ABCD study including household challenges, neglect, and abuse as main categories. Subcategories of ELS data contained parental separation or divorce, criminal household member, household substance abuse, mental Illness in household, mother treated violently in household challenges, emotional neglect, physical neglect in neglect, physical abuse, and sexual abuse in abuse. The items of each subscale were derived from various instruments rated by children themselves or parents: ABCD Youth Family Environment Scale-Family Conflict Subscale Modified from PhenX, ABCD Diagnostic Interview for DSM-5 Traumatic Events, ABCD Family History Assessment, ABCD Parent Demographic Survey, ABCD Children’s Report of Parental Behavioral Inventory, and ABCD Parental Monitoring Survey. We assigned 0 to those who have never had ELS and 1 to those who have at least one type. For three main categories, a child has 1 if at least one subcategory is 1. Missing values were imputed with Bayesian approach and then z-normalization was performed.

#### Brain imaging analysis

Brain imaging included in this study are as follows: structural MRI (sMRI), diffusion MRI (dMRI), resting-state functional MRI (rs-fMRI), and task-based functional MRI. For task-based functional MRI, participants performed three types of tasks including stop signal task (SST), the emotional version of N-back (N-back), and the monetary incentive delay task (MID).

MRI scanning was performed using a 3T scanner (Siemens Prisma, General Electric (GE) 750 and Philips). Image processing and quality assessment for the brain imaging datasets were performed at the ABCD Data Analysis and Informatics Center (DAIC). The ABCD MRI quality assessment consists of three parts: protocol compliance checking, automated quality metrics, manual review of data quality ^18^. Firstly, protocol compliance checking examined whether key imaging parameters matched expected values of a given scanner, such as voxel size or repetition time. Secondly, quality control metrics were obtained for sMRI and fMRI. Mean motion and the number of slices and frames affected by slice dropout by head motion were controlled. Thirdly, the data quality was manually assessed by trained investigators as binary (0 = reject and 1 = accept) considering the image quality and the severity of the artifact (motion, intensity inhomogeneity, white matter underestimation, pial overestimation, magnetic susceptibility artifact). Images that failed to pass the quality assessment were excluded from the analysis.

We acquired multi-shell diffusion MRI from an ABCD study using the following protocol^19^. The ABCD Data Analysis and Informatics Center (DAIC) preprocessed diffusion MRI including distortion and motion correction. B0 distortion and gradient nonlinearity distortion were corrected ^20,21^. We used individualized connectome data to estimate brain imaging phenotypes. Using MRtrix3, we preprocessed diffusion MRI (dMRI), estimated whole-brain white matter tracts, and generated an individualized connectome ^22^. To estimate connectivity, we used streamline counts which represent the fiber connection strength associated with fiber integrity ^23,24^. After decreasing the noise, we performed bias correction with the N4 algorithm of the Advanced Normalization Tools (ANTs) pipeline ^25^. Of the target 20 million streamline counts, we filtered out preliminary tactograms with spherical-deconvolution for a 2:1 ratio. Finally, we generated an 84 × 84 whole-brain connectome matrix for each participant with 10 million streamline counts using T1-based parcellation and segmentation from FreeSurfer. All the computation was carried out by supercomputers at the Argonne Leadership Computing Facility Theta and Texas Advanced Computing Center Stampede 2.

For resting-state functional MRI acquisition, participants completed four 5-minutes resting-state blood oxygen level-dependent scans, with their eyes open and fixated on a crosshair. Participants were also scanned during cognitive tasks. Resting-state functional MRI and task-based functional MRI data were processed and analyzed according to standardized ABCD protocols ^19^. Because of post-processing problems on resting-state and task-based functional MRI data collected on the Philips scanners, all data from the Philips scanner were removed from the current analysis following the ABCD data analysis center’s advice. For functional brain features, we used functional connectivity measures from resting-state fMRI and pairwise correlation coefficients between each region of interest (ROI) from task-based fMRI. The pairwise correlations were examined for ROIs within functionally defined parcellations (i.e., Gordon networks) and subcortical ROIs, and applied Fisher’s r to z-transformation.

#### Deep Neural Network

Firstly, we split the data into discovery and replication sets. Within the discovery set, we trained, optimized, and validated models with stratified 5 fold cross validation. Secondly, we tested the generalizability of the optimized models on the balanced (down-sampled) replication set and then measured model performance with sensitivity, specificity, accuracy, area under the receiver operating characteristic curve (ROC-AUC), and area under the precision recall curve (AUPRC). These metrics were estimated using the pROC package v.1.16.2 in the R programming language and the sklearn package for average precision scores.

The benchmark model contained existing non-imaging variables about suicidality. These variables were also included in all neuroimaging models, since one of our research goals was to test whether neuroimaging data would be useful for suicide prediction.

For multimodal interpretable neuroimaging models for identifying youth suicidality, we used TabNet and further implemented stacking ensemble, GAN-based data augmentation, and dimension reduction methods. TabNet has been developed for tabular data enabling rigorous end-to-end deep learning with built-in interpretability ^26^. This neural network is based on a sequential attention mechanism that gently selects features to infer at each decision step and then accumulates processed information to make final prediction decisions. By using sequential attention mechanisms, selecting sparse features, the model could learn efficiently at each decision-making stage. Therefore, the model with fully related variables shows high performance by taking advantage of related variables. This sparsity could allow for more interpretable decision-making through visualization of variable selection masks ^26^. In this study, given the large feature space of the multimodal brain imaging data, TabNet would enable effective learning of latent representations of the complex data as well as interpretation of the models. Running a single TabNet experiment took around 18 hours on three V100 GPU cards. Hyper-parameter tuning was done by grid search. We changed the width of the attention embedding for each mask, coefficient for feature reusage in the masks, number of steps, learning rate and weight decay. Thus, we used 32-dimension hyper-parameter spaces and found the best hyper-parameter dimension for each dataset.

### Stacking Ensemble Modeling

Stacking ensemble algorithm, widely used ensemble methods along with voting and bagging, is a two-level classification consisting of base classifier level and meta-classifier level^27^. Previous studies showed that a multimodal stacked ensemble algorithm yielded better representation than single modal algorithms ^28,29^. Therefore, we aimed to test the effects of stacking ensembles on multimodal neuroimaging data, and whether stacking ensemble methods take advantage of learning higher-level feature representation.

We generated stacking ensemble models whose output types of base learners are continuous probability values. Trained models of single neuroimaging data with TabNet are base learners. Train data and test data used at the meta-classifier level were concatenated with predicted results from cross-validation and testing in base learners. Logistic regression, xgboost, and random forest were used as meta-learners. The best results from the three meta-learners were reported.

### Data Augmentation

For imbalance between case and controls in suicide prediction that might constrain representation learning, we implemented GAN-based data augmentation approaches through conditional tabular GAN ^30^. Given the fact that youth suicidal ideation and attempts are rare events at the population level, we observed a highly imbalanced distribution between youth at risk for suicidality. Thus, augmented suicidality samples with synthesized data. We generated as many as samples until generated samples showed acceptable CTGAN metrics ^30^: logistic_detection (Logistic Regression classifier to detect whether each row is real or synthetic, the returned score is 1 - ROC-AUC score obtained by the classifier): 0.99, svc_detection(Support Vector Classifier to detect whether each row is real or synthetic): 0.92 in rs-fMRI + sMRI + dMRI dataset and logistic_detection: 0.99, svc_detection: 0.99 in rs-fMRI + SST fMRI + N-back fMRI + MID fMRI dataset. As a result, the augmented discovery set was less imbalanced with a ratio of 1: 2.5.

### Dimension reduction

We utilized TabNet and principal component analysis (PCA) to reduce dimensionality by selecting important features for modeling. For feature selection, we chose the top 50 features (in TabNet) or components (in PCA) in each neuroimaging modality estimated in validation sets. This number was based on our observation that the rate of the increase in explainability slowed for more than 50 features **(Supplementary Figure 1)**. For PCA, the full Singular Value Decomposition method was used ^31^.

## Results

### Demographics

We used data from the ABCD study, a nationwide multisite prospective, longitudinal study. The enrolled samples consisted of 9 and 10-year-old children in the US across 21 sites. Total participants used in this analysis included 837 children with suicidality (i.e., Suicidal ideation or Suicidal attempt) and 5,335 controls **(Table 1)**.

**Table 1.** Sociodemographics of the study participants (n=6,172).

|  | Controls |  | SI |  | SA |  | Statistics | p-value |
| --- | --- | --- | --- | --- | --- | --- | --- | --- |
| N | 5335 |  | 782 |  | 55 |  |  |  |
| Suicidal ideation |  |  |  |  | 51 |  |  |  |
| Age | 119 ± 7.4 |  | 119 ± 7.42 |  | 121 ± 6.08 |  | F = 2.295 | 0.101 |
| Sex |  |  |  |  |  |  |  |  |
| Male | 2704 | 50.68 | 465 | 59.46 | 36 | 65.45 | Chisq = 25.123 | p<.001 |
| Female | 2631 | 49.32 | 317 | 40.54 | 19 | 34.55 |  |  |
| Race / Ethnicity |  |  |  |  |  |  |  |  |
| White | 2994 | 56.12 | 426 | 54.48 | 20 | 36.36 | Chisq = 32.56 | p<.001 |
| Black | 482 | 9.03 | 69 | 8.82 | 13 | 23.64 |  |  |
| Hispanic | 1183 | 22.17 | 157 | 20.08 | 18 | 32.73 |  |  |
| Asian | 120 | 2.25 | 20 | 2.56 | 1 | 1.82 |  |  |
| Other | 551 | 10.33 | 110 | 14.07 | 3 | 5.45 |  |  |
| Parents married | 3858 | 72.31 | 517 | 66.11 | 26 | 47.27 | Chisq = 28.489 | p<.001 |
| Income level | 7.42 ± 2.26 |  | 7.22 ± 2.29 |  | 5.66 ± 2.6 |  | F = 18.551 | p<.05 |

### Identification of children with suicidal ideation using neuroimaging data

Our neuroimaging models for classification of suicidal ideation showed varied performances ranging from test ROC-AUCs of 48.26 to 61.22 (AURPCs: 47.85—60.06; **Table 2**). Among the models trained on neuroimaging features only, the structural MRI, the diffusion MRI and the resting-state fMRI models (ROC-AUCs: 61.22—57.18, AURPCs: 52.74—60.06) provided higher performances than task fMRI models (ROC-AUCs: 48.26—53.42, AURPCs: 47.85—50.87), which exceeded at chance level. All the neuroimaging models fell short of the classification performance of the psychosocial model, which yielded a moderate accuracy in classifying children with suicidal ideation and controls (ROC-AUC = 74.16, AUPRC: 70.18).

**Table 2.** Classification performances of psychosocial, imaging-based, and combined deep neural networks identifying suicidal ideation. Deep neural network (TabNet) and additional methods for dimensionality reduction (feature selection, PCA) and data augmentation (CTGAN) were tested.

| Model | Replication set |  |  |  |  |
| --- | --- | --- | --- | --- | --- |
|  | ROC-AUC | AUPRC | Accuracy | Sensitivity | Specificity |
| **Psychosocial model** | 74.16 | 70.18 | 70.66 | 72.78 | 68.55 |
| **Structural MRI** |  |  |  |  |  |
| Neuroimaging model | 57.18 | 52.74 | 57.53 | 43.81 | 70.18 |
| Combined model | 71.44 | 67.95 | 67.82 | 84.18 | 51.57 |
| Neural net based feature selected | 72.55 <sup>†</sup> | 69.05 | 69.72 | 75.32 | 64.15 |
| PCA based dimension reduced | 67.44 | 63.18 | 65.93 | 71.52 | 60.38 |
| GAN based data augmented | 69.74 | 65.08 | 67.51 | 78.48 | 56.60 |
| **Diffusion MRI** |  |  |  |  |  |
| Neuroimaging model | 58.15 | 53.48 | 58.9 | 64.76 | 53.51 |
| Combined model | 69.54 | 64.79 | 66.88 | 74.68 | 59.12 |
| Neural net based feature selected | **74.82** * | 70.06 | 72.24 | 75.95 | 68.55 |
| PCA based dimension reduced | 69.1 | 65.31 | 66.56 | 75.95 | 57.23 |
| GAN based data augmented | 68.74 | 64.09 | 65.93 | 73.42 | 58.49 |
| **Resting state fMRI** |  |  |  |  |  |
| Neuroimaging model | 61.22 | 60.06 | 59.82 | 82.86 | 38.6 |
| Combined model | 73.74 | 67.58 | 70.34 | 86.07 | 54.71 |
| Neural net based feature selected | 73.43 | 70.86 * | 69.72 | 72.78 | 66.67 |
| PCA based dimension reduced | 69.95 | 62.02 | 67.51 | 81.01 | 54.09 |
| GAN based data augmented | 71.26 | 66.92 | 67.51 | 81.65 | 53.46 |
| **Stop signal task fMRI** |  |  |  |  |  |
| Neuroimaging model | 53.42 | 50.87 | 54.79 | 55.24 | 54.39 |
| Combined model | 71.37 | 67.6 | 67.51 | 65.79 | 69.11 |
| Neural net based feature selected | 72.52<sup>†</sup> | 68.41 | 67.93 | 62.28 | 73.17 |
| PCA based dimension reduced | 66.61 | 61.22 | 66.24 | 65.79 | 66.67 |
| GAN based data augmented | 70.24 | 65.05 | 66.24 | 71.05 | 61.79 |
| **N-back task fMRI** |  |  |  |  |  |
| Neuroimaging model | 50.43 | 47.85 | 53.88 | 50.48 | 57.02 |
| Combined model | 72.69 | 66.73 | 70.04 | 81.58 | 59.35 |
| Neural net based feature selected | 72.09 | 68.71 | 68.78 | 75.44 | 62.60 |
| PCA based dimension reduced | 68.29 | 61.99 | 65.82 | 54.39 | 76.42 |
| GAN based data augmented | 68.78 | 62.29 | 66.67 | 85.09 | 49.59 |
| **Monetary incentive delay task fMRI** |  |  |  |  |  |
| Neuroimaging model | 48.26 | 49.47 | 54.79 | 64.76 | 45.61 |
| Combined model | 71.22 | 66.19 | 65.82 | 65.79 | 65.85 |
| Neural net based feature selected | 72.28<sup>†</sup> | 68.89 | 70.04 | 71.93 | 68.29 |
| PCA based dimension reduced | 64.58 | 60.86 | 64.14 | 62.28 | 65.85 |
| GAN based data augmented | 68.23 | 63.39 | 66.67 | 73.68 | 60.16 |

Note: Combined models included imaging derived and psychosocial features. \* indicates higher performance than the benchmark result in TabNet and † indicates higher performances than TabNet results in the same dataset.

### Combining psychosocial and imaging-based data showed improvements in identifying children with suicidality

When combining behavioral and brain imaging-derived phenotypes, models showed moderately accurate performance in classifying children with suicidal ideation, ranging from ROC-AUC 69.54% to ROC-AUC 74.16% (AUPRC: 64.79—67.85; **Table 2**). Moreover, to address problems brought from the large feature space of multimodal neuroimaging data, we implemented dimension reduction and data augmentation strategies to further optimize the suicide prediction models. We found that deep neural net-based feature selection led to marginal improvement of ROC-AUC in the combined models of structural MRI, diffusion MRI, SST fMRI, and MID fMRI. Of note, the diffusion MRI model outperformed the benchmark model by a small margin (diffusion MRI: 74.82 ROC-AUC, 70 AUPRC).

### Integration of multimodal neuroimaging data

We further investigated whether fusing psychosocial and multimodal neuroimaging data could improve the identification of suicidal ideation. While most multimodal neuroimaging models failed to show better classification than the benchmark model, two models (i.e., dMRI + rs-fMRI, rs-fMRI + sMRI + dMRI) showed comparable ROC-AUCs to that of the benchmark model **(Table 3)**. These models were based on feature selection and ensemble methods.

**Table 3.** Classification performances (ROC AUC) of psychosocial and multimodal neuroimaging data for identification of suicide ideation on balanced replication datasets.

|  | Model | TabNet | TabNet - | Ensemble |
| --- | --- | --- | --- | --- |
|  |  |  | Feature selection |  |
| Morphometry and connectivity | sMRI + dMRI | 73.81 ‡ | 72.3 | 73.91<sup>†</sup> |
|  | sMRI + rs-fMRI | 72.43 | 72.71<sup>†</sup> | 73.77<sup>†</sup> |
|  | dMRI + rs-fMRI | 72.86 | 73.28 | **74.7\*** |
|  | rs-fMRI + sMRI + dMRI | 68.47 | **74.43 \*** | **74.41\*** |
| Functional connectivity and activation | rs-fMRI + SST fMRI | 73.58 ‡ | 72.23 | 54.97 |
|  | rs-fMRI + Nback fMRI | 73.82 ‡ | 72.31 | 56.22 |
|  | SST fMRI + Nback fMRI | 71 | 72.48<sup>†</sup> | 56.71 |
|  | SST fMRI + Nback fMRI + MID fMRI | 71 | 72.74<sup>†</sup> | 53.94 |
|  | rs-fMRI + SST fMRI+ Nback fMRI + MID fMRI | 70.43 | 69.91 | 56.51 |
| All modalities | sMRI + dMRI + rs-fMRI + SST fMRI + Nback fMRI + MID fMRI | 65.9 | 69.76<sup>†</sup> | 54.95 |

\* represents higher performances than the psychosocial model with ROC AUC of 74.16. † denotes higher performances than TabNet results in the same dataset. ‡ denotes higher performances than the combined model trained on single neuroimaging data.

Also, the combined model of sMRI and dMRI, which represents brain morphology and structural connectivity of white matter, offered higher ROC-AUC compared with the single modal performances of sMRI and dMRI, respectively. We also found that integrating functional MRIs slightly exceeded the performance of each single modality model: e.g., rs-fMRI + SST fMRI: 73.58 ROC-AUC, rs-fMRI + N-back fMRI: 73.82 ROC-AUC.

Compared with the same multimodal models trained on original features, the feature selected models provided slightly higher performances. In ensemble models, while the connectivity models (i.e., sMRI, dMRI, rs-fMRI) showed slight improvements in test ROC-AUCs, the models trained on task fMRI-derived activation estimates showed a sharp decrease to 53.94—56.71 ROC-AUCs.

### Model Interpretation

Across modalities, the feature importance plots of our models included brain derived features related to anterior cingulate cortex, temporal gyrus, and precentral gyrus **(Figure 1)**. Common psychological features were observed including Internalizing-externalizing comorbidity, Prodromal psychosis, Anxiety, Depression, and Family conflict. These multimodal features showed patterns in predicting the risk for suicidal ideation across modalities: top 2∼17% features accounted for 90% performances. These important features consisted of small numbers of psychological features (9∼17%) and large amounts of neuroimaging derived features (83∼90%). Complete feature importance details are in **Supplementary Figure 2**.

**Figure 1.**
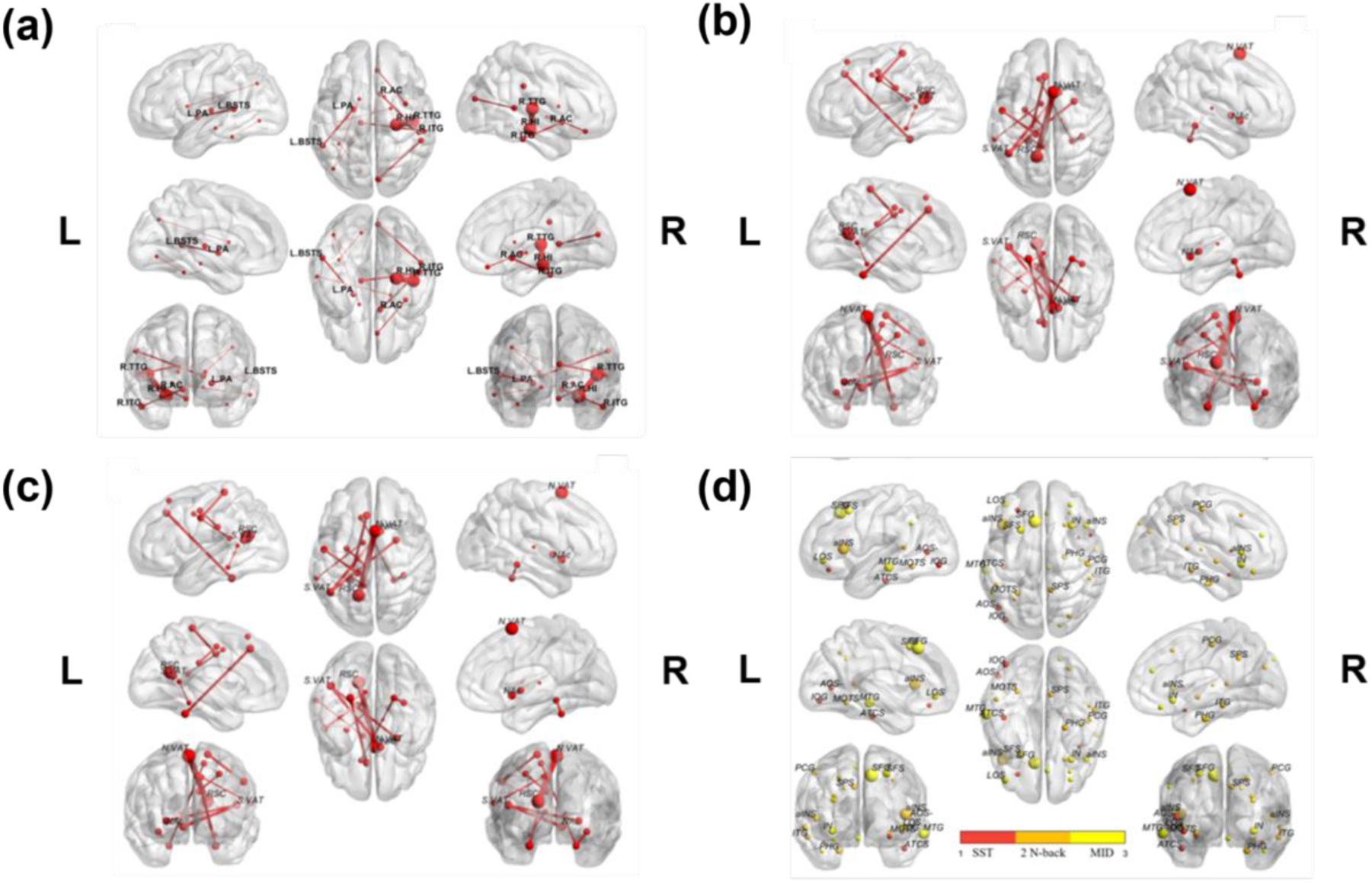
Important neuroimaging features contributing to the identification of suicidal ideation in combined models with selected features. A. dMRI-derived White Matter Connectivity. B. Structure MRI-Derived Morphometry. C. RS-FMRI-Derived Functional Connectivity. D. Task FMRI-Derived Activation Estimates (Stop signal task, emotional N-back task, monetary incentive delay task). The size of the node and edge represents the relative importance. **dMRI-derived White Matter Connectivity** L.BSTS: Left Banks of Superior Temporal Sulcus, L.PA: Left-Pallidum. R.AC: Right-Accumbens-area, R.HI: Right-Hippocampus, R.ITG: gray matter of right inferior temporal gyrus, R.TTG: gray matter of right transverse temporal gyrus./ **Structure MRI-Derived Morphometry** caudalMFG: caudal middle frontal, frontal pole: frontal pole, fusiform-G: fusiform gyrus, Insula: superior segment of_the circular sulcus of the_insula, lingual-gyrus: lingual-gyrus, occipital-pole: occipital pole, rostral-ACC: rostral anterior cingulate, subcallosal-G: subcallosal gyrus/ **RS-FMRI-Derived Functional Connectivity** N.VAT: Network ventral attention, NAc: nucleus accumbens, RSC: retrosplenial temporal cortex, S.VAT: Subcort ventral attention, / **Task FMRI-Derived Activation Estimates** aINS: vertical ramus of the anterior segment of the lateral sulcus, aINS: vertical ramus of the anterior segment of the lateral sulcus, AOS: anterior occipital sulcus and preoccipital notch, ATCS: anterior transverse collateral sulcus, IN: superior segment of the circular sulcus of the insula, IOG: inferior occipital gyrus and sulcus, LOS: lateral orbital sulcus, MTG: middle temporal gyrus, PCG: postcentral gyrus, PHG: parahippocampal gyrus, SFG: superior frontal gyrus, SFS: superior frontal sulcus, SPS: subparietal sulcus

**Figure 2.**
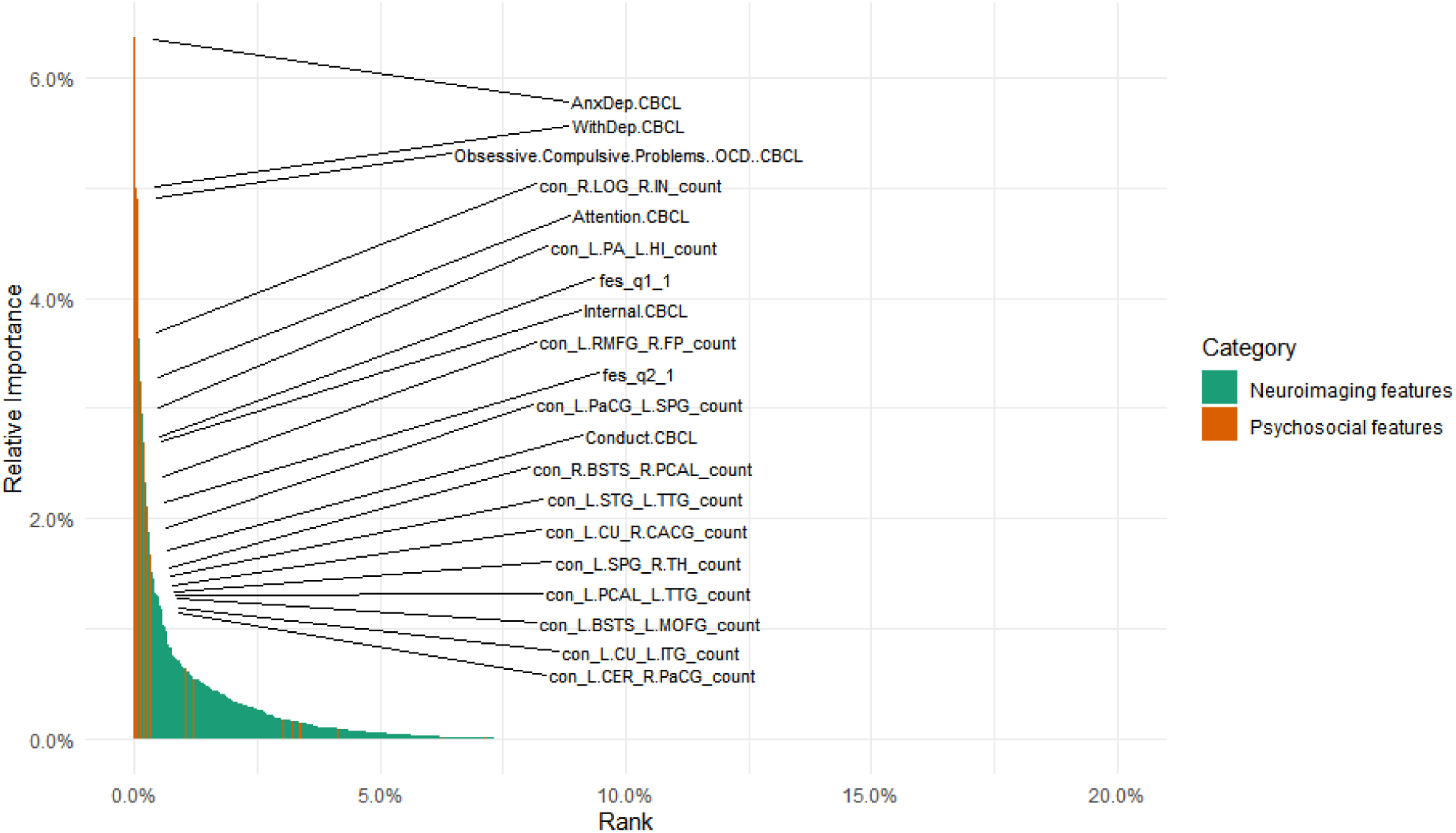
Feature importance plot of diffusion MRI combined model. Important 20 features are indicated. Top 5% features (N=180) exceed 95% of cumulative importance. AnxDep.CBCL: Anxious/depressed behavior, Attention.CBCL: Attention problems, con_R.LOG_R.IN_count: connectivity between right lateral occipital gyrus and right insula, con_L.PA_L.HI_count: connectivity between left pallidum and left hippocampus, con_L.RMFG_R.FP_count: connectivity between left rostral middle frontal gyrus and right frontal pole, con_L.PaCG_L.SPG_count: connectivity between left parahippocampal gyrus and right subparietal gyrus, con_R.BSTS_R.PCAL_count: connectivity between right bankssts and right pericalcarine, con_L.STG_L.TTG_count: connectivity between left superior temporal gyrus and left transverse temporal gyrus,, con_L.CU_R.CACG_count: connectivity between left cuneus and right caudal anterior cingulate, con_L.SPG_R.TH_count: connectivity between left superior parietal gyrus and right thalamus, con_L.PCAL_L.TTG_count: connectivity between left pericalcarine and right transverse temporal gyrus, con_L.BSTS_L.MOFG_count: connectivity between left bankssts and right medial orbitofrontal gyrus, con_L.CU_L.ITG_count: connectivity between left cuneus and gray matter of left inferior temporal gyrus, con_L.CER_R.PaCG_count: connectivity between left Cerebellum-Cortex and right paracentral gyrus, Conduct.CBCL: Conduct problems, conflict_openly angry, fes_q1_1: Family_frequent fights, fes_q2_1: Family, Internal.CBCL: Internalizing behavior, Obsessive.Compulsive.Problems..OCD..CBCL: OCD problems, WithDep.CBCL: Withdrawn/depressed behavior

## Discussion

We tested deep neural networks trained on the largest youth brain multimodal MRI data, as well as parent-reported psychosocial questionnaires, to predict youth with suicidal thoughts. Our deep neural networks showed both feasibility and limitations of the brain-based risk prediction of youth suicidality. Throughout our systematic model comparison, the one trained on white matter connectivity and psychological data performed the best. Models trained on multimodal brain imaging-derived data alone, including white matter connectivity, morphometry, functional connectivity, and task reactivity, showed above-chance performance yet were poorer compared with the baseline psychological model. Model interpretation showed the cortical regions and connections of which structural or functional metrics contributed to the prediction of youth suicidality, including the inferior frontal, temporal, precentral gyri, insula, and anterior cingulate cortex. These results from large, multi-site, epidemiological samples show the feasibility of the data-driven computational learning approach to multimodal brain MRI for prediction, as well as scientific discovery of youth suicidality.

The brain alone models showed statistically meaningful classification performance with the maximum ROC-AUC of 61.22 in resting-state fMRI-functional connectivity and similarly in structural and diffusion MRI-derived estimates. Prior studies with small sample sizes (e.g., N < 160) in adults report moderate accuracy in predicting suicidal ideation or attempts based on structural and functional brain imaging (with ROC-AUC ranging from 0.72 to 0.94; some results from only cross-validation but not from held-out test set)^6,7,32,33^. Conversely, literature in children has been extremely rare. To our knowledge, this is the first study reporting the machine learning application to neuroimaging data to identify youth with suicidality.

Our neural networks permit delineating the brain correlates of youth suicidality. Our integrative models combining the psychosocial and white matter connectivity showed a slight increase in model performance (with a 1% ROC-AUC) compared with the baseline psychosocial model. Important features in this model include white matter connectivity, morphometry, and functional connectivity within the distributed brain network, primarily the neocortex, including the inferior frontal gyrus, insula, and anterior cingulate cortex. The inferior frontal gyrus plays a role in emotional regulation, sustained attention, and language processing ^34^, of which abnormalities were associated with suicidality in adolescents ^35^. The insula and anterior cingulate cortex, as part of the salience network, are involved in the detection and integration of emotional and sensory stimuli ^36^. These features selected by the deep neural network with the sequential attention mechanism, are largely overlapped with those selected in the linear mass-univariate analysis in a prior study^37^ (note that this prior study did not include diffusion MRI). Despite neither the mass-univariate nor machine learning approach can present the causal brain circuitry underlying suicide, the brain circuit correlates found in this study may help future research in youth suicidality.

Considering the previous univariate statistical analysis of the same data reporting the low classification accuracy (with area under precision-recall less than 0.10) for youth suicidality using brain imaging (cf. without diffusion data)^37^, our results of the maximum ROC-AUC of 0.75 (area under precision-recall of 0.70) in held-out test samples show the importance of nonlinear multimodal modeling with machine learning in individualized prediction. Then, a more highly parameterized, end-to-end learning model trained on 3D and 4D brain MRI would improve accuracy to the level of practicality. Despite that a recent study shows a poorer scalability of the end-to-end deep learning models than linear models with the size of 10,000 samples ^38^, which was based on the tasks to predict common biological phenotypes (e.g., age and sex), we believe it is necessary to test the very scalability of this approach in the task of youth suicidality prediction. Moreover, even if the brain imaging itself contributes to the prediction with a limited degree, integrating the multi-modal data, not limited to the brain imaging, but extending to genetics, microbiome, life-log data, SNS and speech data may help us build a better predictive model with the practical utility in clinical or school settings.

Our deep neural networks trained on psychological and multimodal neuroimaging data may reflect the interplay between psychological variables and brain circuits related to risk for youth suicidality. This is based on our observations of model interpretation across modalities: About 20% of features accounted for more than a 90% variance. Of those, besides several psychological features (e.g., internalizing symptoms) the multimodal brain phenotypes constitute the majority. Although the performance boost owing to the brain data was limited with up to a 1% ROC-AUC increase, we find this observation may reflect the interaction between the internalizing symptoms and the distributed brain system contributing to youth suicidality.

Our models include common psychosocial features as important attributes (e.g., internalizing and depressive behaviors, prodromal psychosis, and family conflict). This result is consistent with previous suicidality literature and recent findings from the ABCD study ^2,10,39^.

We tested two different methods of dimensionality reduction to overcome the sample complexity problem, i.e., the large feature space compared with the sample size. Overall, performance improvements were trivial. TabNet-based feature selection significantly improved performance compared to the models trained on original features, while PCA showed no improvements. Since TabNet’s sequential attention mechanism accounts for the nonlinearity of the relationships among the features, which PCA cannot do so, this result implies the importance of non-linear modeling of the brain data in predicting suicidality in children.

Our stacking ensemble approach showed no benefits of integrating multi-modal brain imaging data. We believe that the stacking approach may have failed to account for complementary characteristics across the modalities. For better multi-modal integration, a principled approach that can utilize multitude representations as well as the brain network organization might be tested in the future. A recent breakthrough in computational chemistry may be a good example. A graph neural network accurately predicts protein folding structures and interactions by learning rich representations about proteins from multimodal and multidimensional data^40^.

Likewise, in our experiment, performance increases by GAN-based data augmentation were not meaningful in the replication set (while it significantly improved performance in the validation set). This may be due to different distributions between the discovery and replication set^41^. In the future, since the benefits of the dimensionality reduction were observed in this study (e.g., TabNet’s sequential attention mechanism), it will be interesting to test the data augmentation method in reduced numbers of the brain features.

There are some limitations to be considered in future studies. Firstly, suicidality (which includes suicidal ideation and attempt) does not always lead to suicidal commitment ^42^. This might result from the difficulty not only of collecting data on who committed suicide but also of obtaining sufficient samples of suicide attempts from prepubertal children. Secondly, this study used a cross-sectional design. This limits us to testing the causal relationship between brain data and suicidality, or predictability of future outcomes. Given that the brain organizations of prepubertal children mature throughout adolescence, future research should test prospective predictions of suicidal risk using longitudinal data.

In sum, this study demonstrates the potential utility of a deep neural network approach using both psychosocial variables and neuroimaging to identify prepubertal children at risk for suicide in large, representative, and multi-site samples.

## Data Availability

The ABCD data can be accessed via the NIMH Data Archive (https://nda.nih.gov/). Our codes used in this study are freely accessible: for the Tabnet implementation (https://colab.research.google.com/drive/1y5I89AxrfGYAlJmW2jnYk42OOlDfP8Zz?usp=sharing) and for the ensemble modeling (https://colab.research.google.com/drive/1lR1Y5BBEuGtxQfDDYZ2RL9EtL__h_Jxf?usp=sharing)

## Acknowledgments

This work was supported by the New Faculty Startup Fund from Seoul National University (Cha); the BK21 FOUR Program (5199990314123) through the National Research Foundation of Korea (Cha and Ahn); National IT Promotion Agency GPU award (Cha); Intel PRTI award (Cha).

## Supplementary Materials

**Supplementary Table 1.** Classification performances in predicting youth suicidality attempt. Deep neural networks (TabNet) and additional methods of dimensionality reduction and data augmentation were tested.

| Model | TabNet | TabNet - Feature selection | TabNet - PCA | TabNet - CTGAN |
| --- | --- | --- | --- | --- |
| Psychosocial | 83.33<sup>‡</sup> | 69.44 | - | 69.44 |
| Structural MRI | 60.68 | 75<sup>†</sup> | 75<sup>†</sup> | 75<sup>†</sup> |
| Diffusion MRI | 72.22<sup>‡</sup> | 79.17<sup>†</sup> | 72.2 | 79.17<sup>†</sup> |
| Resting state fMRI | 80.56<sup>‡</sup> | 80.56 | 84.72 * | 80.56 |
| Stop signal task fMRI | 78.1<sup>‡</sup> | 76.91 | 64.29 | 76.91 |
| N-back task fMRI | 69.05 | 78.57<sup>†</sup> | 76.19<sup>†</sup> | 78.57<sup>†</sup> |
| Monetary incentive delay task fMRI | 54.76 | 54.76 | 78.57 | 54.76 |

Note: All single modality models except psychosocial model included psychosocial by default. \* denotes higher performance than the benchmark result in TabNet and † denotes higher performances than TabNet results in the same dataset. ‡ denotes higher performances than the results in suicidality ideation using TabNet.

On the single modal neuroimaging data, the neural networks, TabNet showed a wider range of accurate performances in classifying children with suicidal attempts, ranging from ROC-AUC 54.76% to ROC-AUC 83.33%.

When we apply feature selection to TabNet, in structural and diffusion MRI, additional methods including feature selection, PCA and CTGAN showed higher performance than just TabNet model. Although there isn’t any higher result than benchmark in both MRI data, we found that dimensionality reduction explains suicidality ideation and attempt better than the other ways.

**Supplementary Table 2.** Summed contribution of top 50 important features of the combined models to identify children at risk for suicidal ideation using TabNet.

| Model | Psychosocial variable | Neuroimaging variables |
| --- | --- | --- |
| Structural MRI | 0.343 | 0.334 |
| Diffusion MRI | 0.296 | **0.359** |
| Resting state fMRI | 0.585 | 0.133 |
| Stop signal task fMRI | 0.382 | 0.343 |
| N-back fMRI | 0.435 | 0.245 |
| Monetary incentive delay fMRI | 0.456 | 0.252 |

**Supplementary Table 3.** Summed contribution of top 50 important features of suicidal attempt prediction models using TabNet

| Model | Psychosocial variable | Neuroimaging variables |
| --- | --- | --- |
| Structural MRI | 0.218 | 0.307 |
| Diffusion MRI | **0.329** | 0.143 |
| Resting state fMRI | 0.129 | 0.411 |
| Stop signal task fMRI | 0.0966 | 0.453 |
| N-back fMRI | 0.105 | 0.396 |
| Monetary incentive delay fMRI | 0.0321 | 0.414 |

In the suicide attempt prediction task, we observed that most neuroimaging variables outweighed psychosocial variables in several neuroimaging models (structural MRI, resting-state fMRI, stop signal task fMRI, N-back task fMRI and monetary incentive delay task fMRI models). However, only diffusion MRI derived features outperformed psychosocial variables among the top 50 features.

**Supplementary Table 4.** Top 10 features predictive of suicidal ideation. The models were trained on brain imaging-derived phenotypes and psychosocial data

| Psychosocial |  | Structural MRI |  | Diffusion MRI |  | Resting state fMRI |  |
| --- | --- | --- | --- | --- | --- | --- | --- |
| Feature | Importance | Feature | Importance | Feature | Importance | Feature | Importance |
| Internalizing-externalizing comorbidity | 15.15 | Anxiety / Depression | 10.18 | Internalizing-externalizing comorbidity | 12.8 | Depression | 6.02 |
| Prodromal psychosis_frequency | 7.37 | Prodromal psychosis_distress | 6 | Prodromal psychosis_frequency | 7 | Prodromal psychosis_frequency | 5.33 |
| Prodromal psychosis_distress | 6.68 | Family temper | 3.71 | Prodromal psychosis_distress | 6.05 | Withdrawn / Depressed | 3.18 |
| Family conflict | 5.53 | Family anger | 3.22 | Family anger | 3.45 | Obsessive-Compulsive Disorder(OCD) | 2.56 |
| Anxiety / Depression | 3.38 | Oppositional Defiant Disorder | 3.07 | Behavioral inhibition system | 3.23 | Weight | 2.44 |
| Depression | 3.01 | Externalizing disorder | 3.05 | Rash tendency in extreme | 2.72 | Internal comorbidity | 2.39 |
| Family angry | 2.65 | ADHD | 2.68 | Connectivity_Right-Hippocampus_Gray matter of right transverse temporal gyrus | 2.65 | Social Problems | 2.11 |
| Family peace | 2.57 | Sex | 2.54 | Depression | 2.61 | Married | 2.1 |
| Income | 2.35 | Internal comorbidity | 2.52 | Aggressive behavior | 2.56 | Family Angry | 1.97 |
| Internal comorbidity | 2.2 | Total Problem | 2.46 | Family temper | 2.12 | Connectivity_retrosplenialtemporal_dorsalattention | 1.92 |
|  |  | Stop signal task fMRI |  | N-back task fMRI |  | Monetary incentive delayed task fMRI |  |
|  |  | Feature | Importance | Feature | Importance | Feature | Importance |
|  |  | Depression | 9.19 | Prodromal psychosis_frequency | 10.77 | Prodromal psychosis_frequency | 9.75 |
|  |  | Prodromal psychosis_frequency | 6.41 | Anxiety / Depression | 6.12 | Aggressive Behavior | 7.35 |
|  |  | Externalizing disorder | 5.74 | Externalizing disorder | 5.37 | Prodromal psychosis_distress | 7.04 |
|  |  | Prodromal psychosis_distress | 5.55 | Aggressive behavior | 3.71 | Obsessive-Compulsive Disorder(OCD) | 6.4 |
|  |  | Internal comorbidity | 5.3 | Rule breaking behavior | 3.4 | Depression | 5.51 |
|  |  | Family anger | 3.27 | Depression | 3.1 | Internalizing-externalizing comorbidity | 4.76 |
|  |  | Anxiety / Depression | 2.75 | Internalizing-externalizing comorbidity | 2.8 | superior frontal gyrus (L) | 2.74 |
|  |  | Internalizing-externalizing comorbidity | 2.53 | Family conflict | 2.47 | Externalizing disorder | 2.64 |
|  |  | Obsessive-Compulsive Disorder(OCD) | 2.39 | Conduct disorder | 2.43 | Family temper | 2.16 |
|  |  | Race | 1.85 | Obsessive-Compulsive Disorder(OCD) | 2.41 | middle temporal gyrus (L) | 9.75 |

**Supplementary Figure 1.**
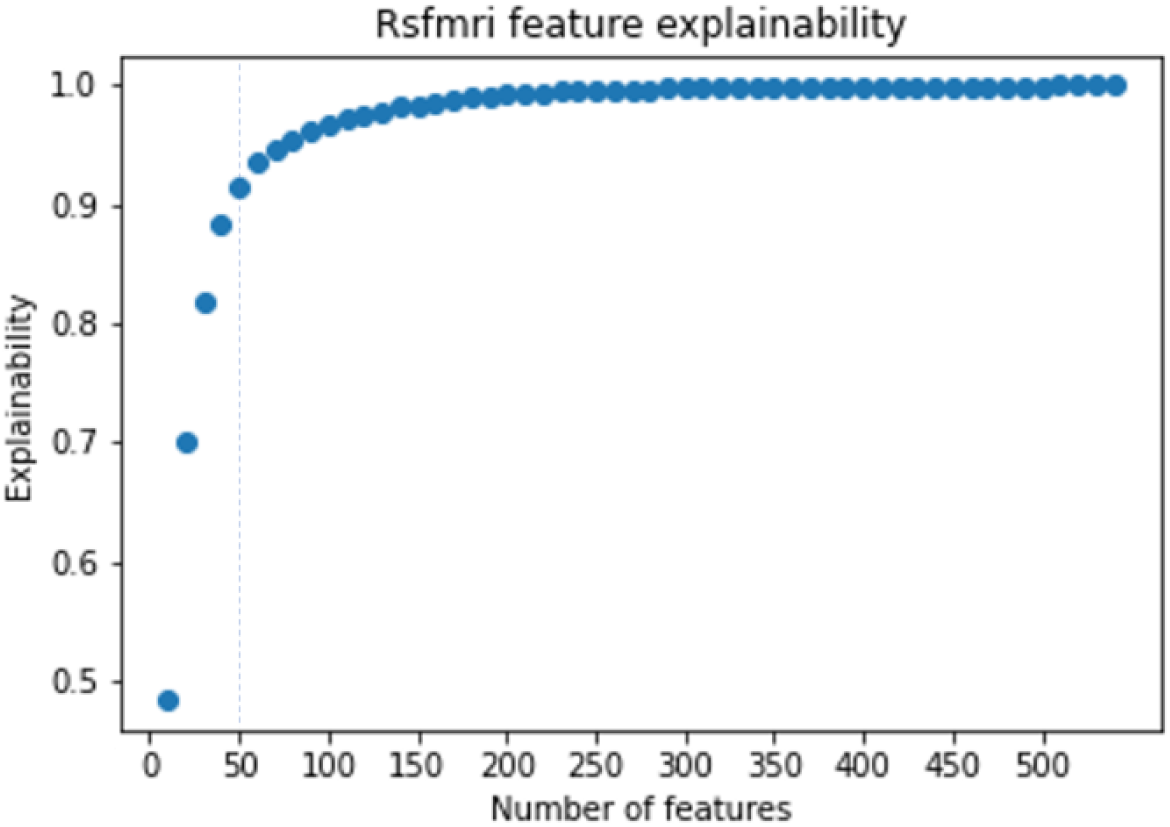
Explainability of the resting-state fMRI combined model as an increase of the number of features. Top 50 features contain explainability of 91.6%.

**Supplementary Figure 2.**
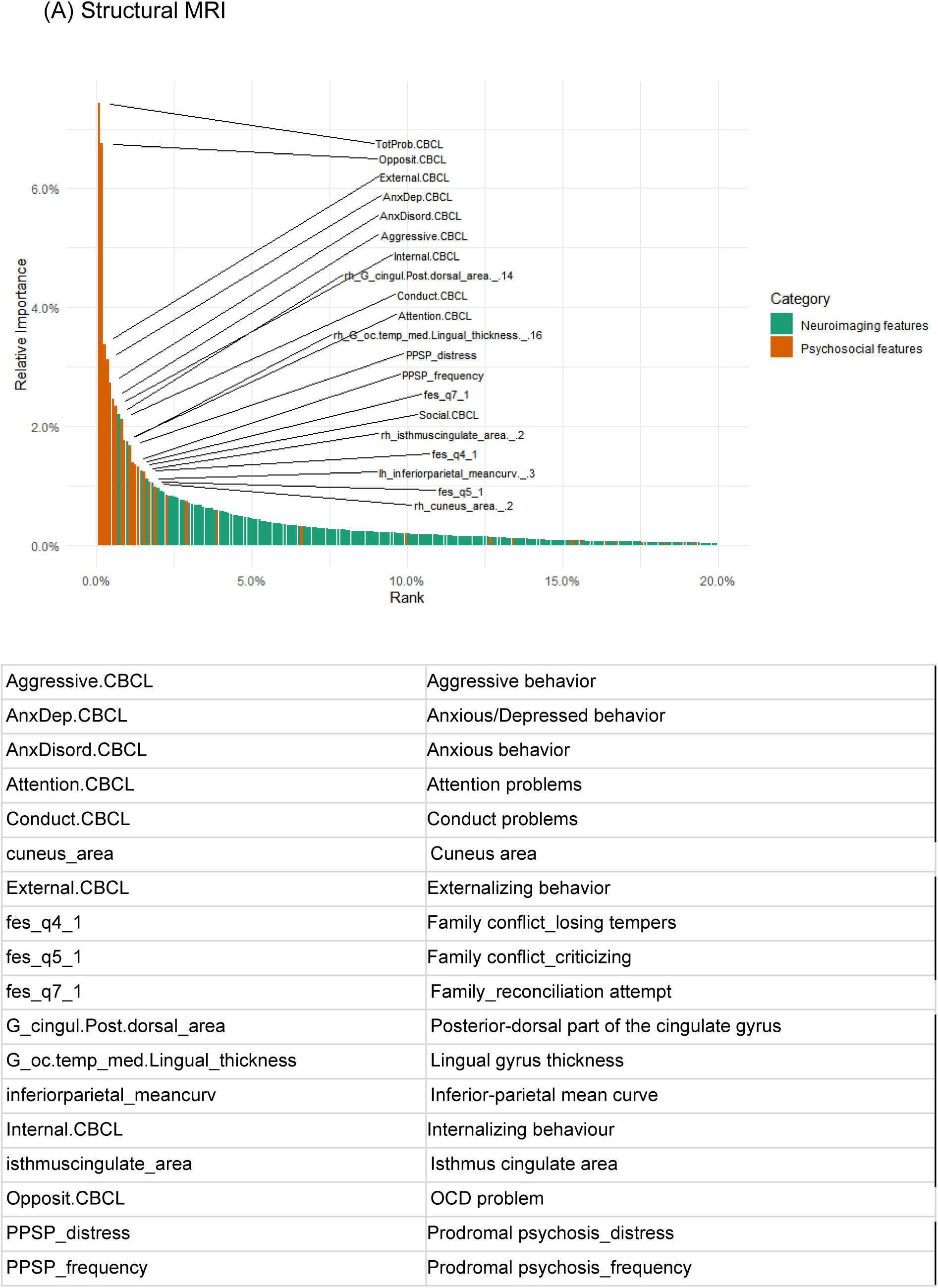
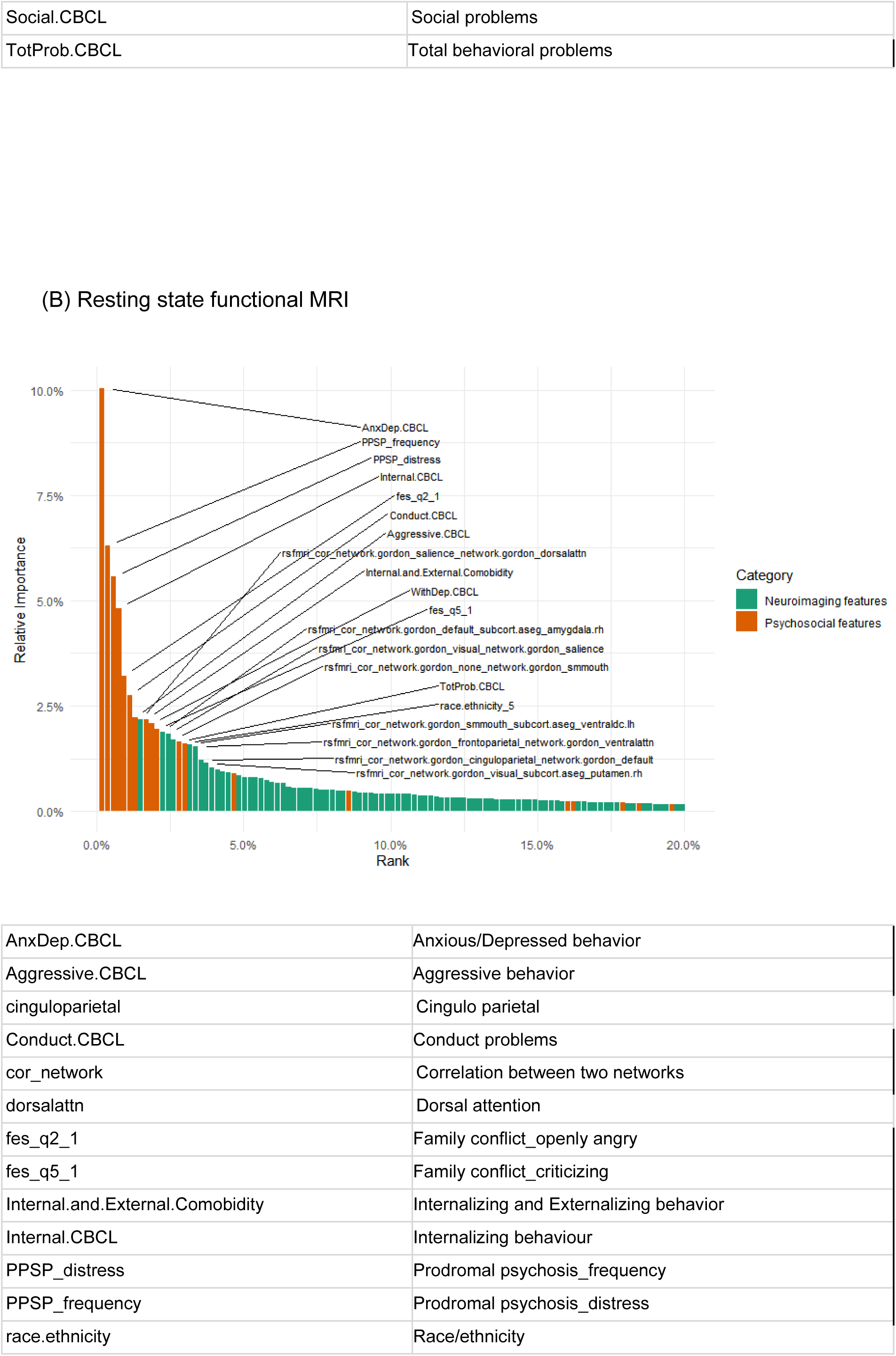
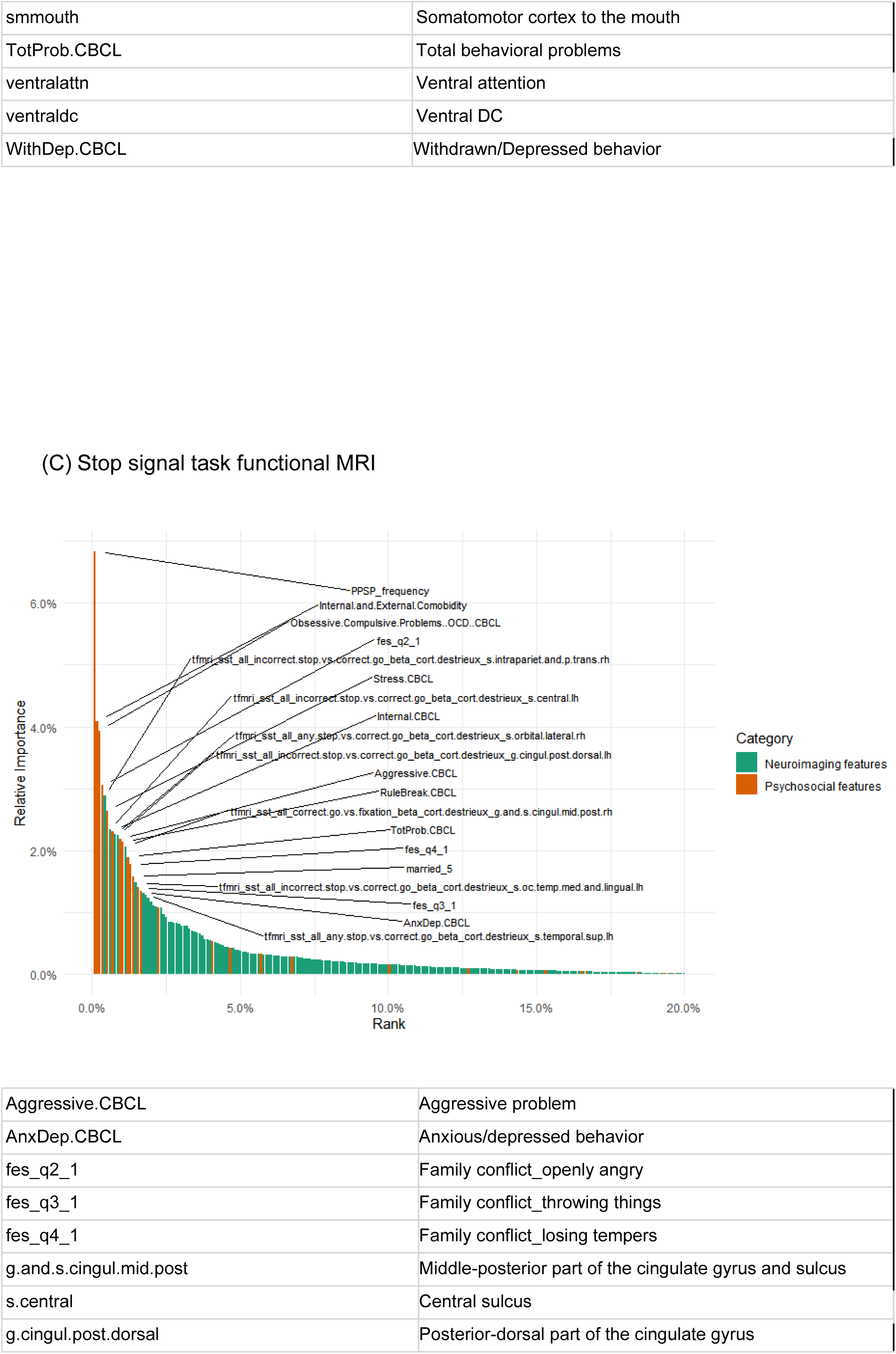
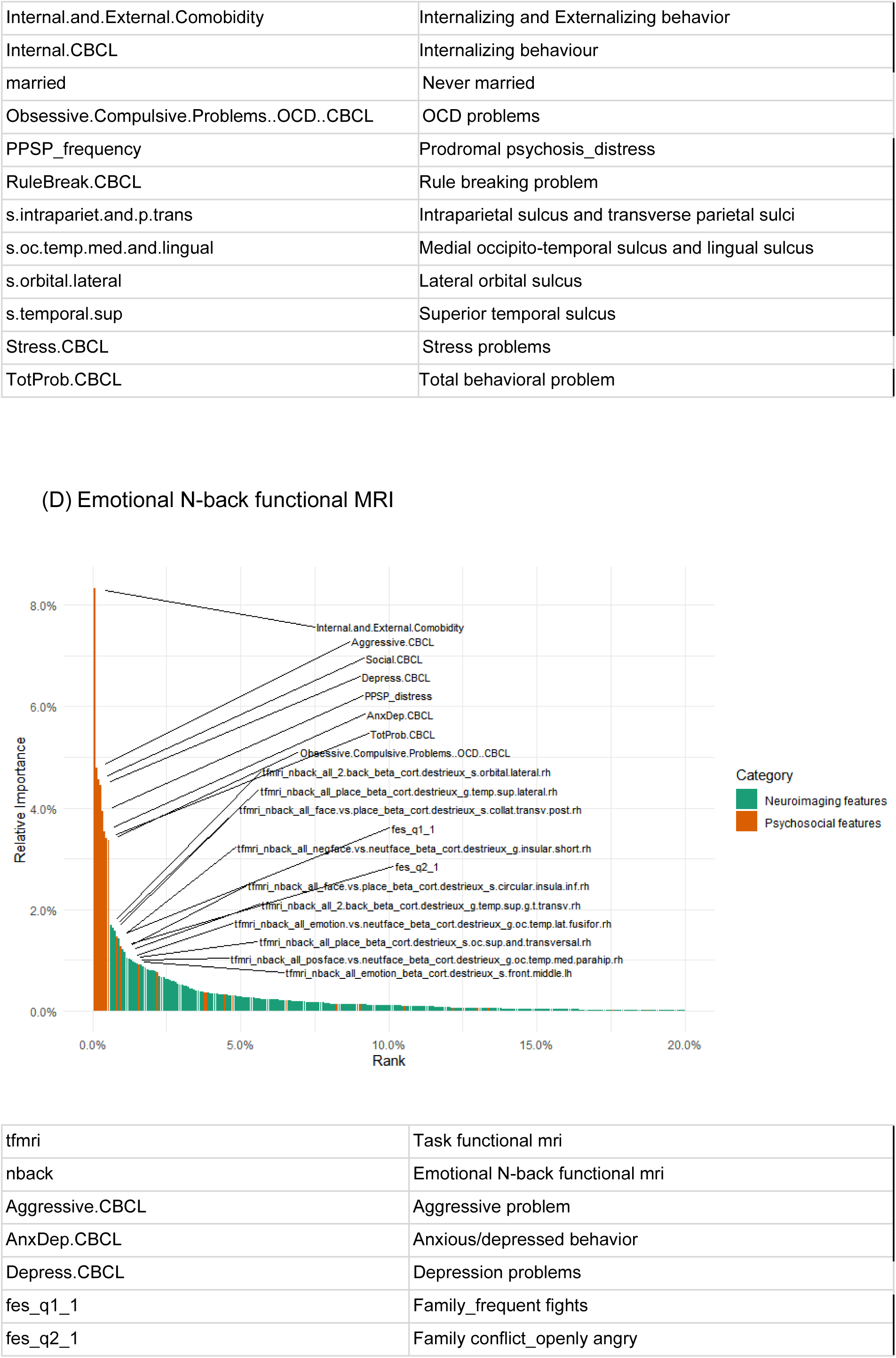
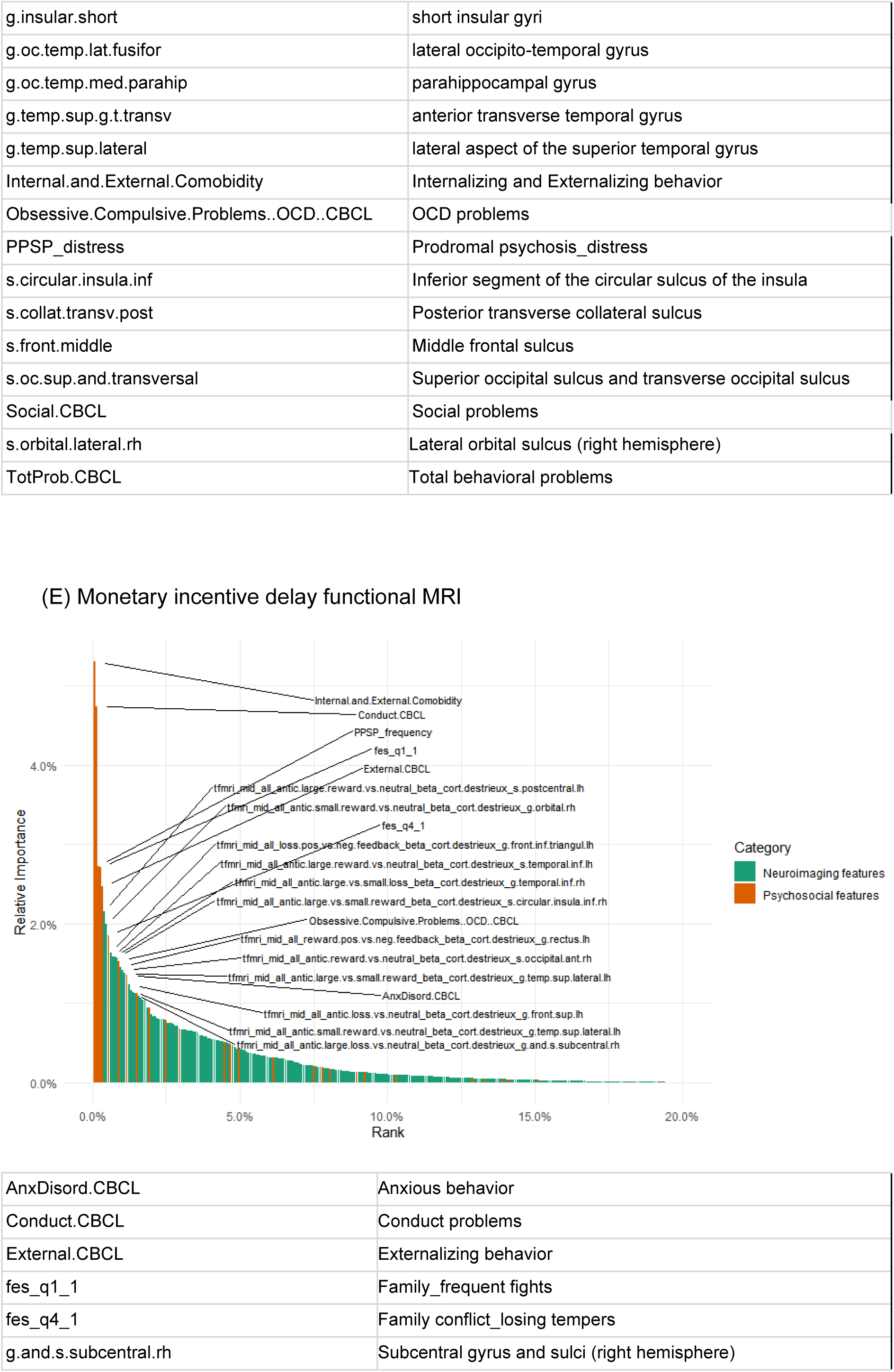
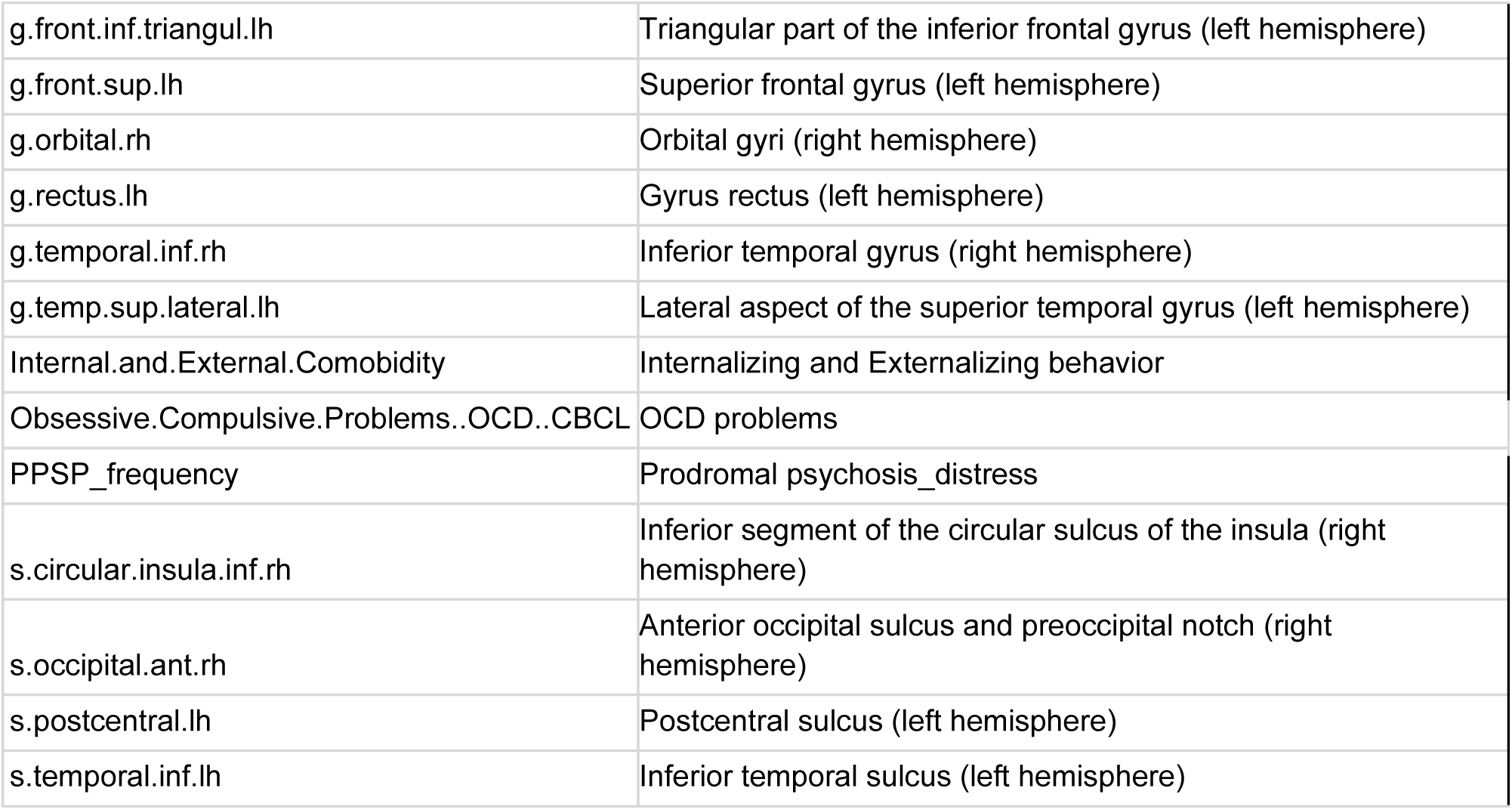
Feature importance plots of combined models. Top 20 features are denoted.

